# College Students’ COVID-19 Vaccine Beliefs and Intentions: Implications for interventions

**DOI:** 10.1101/2021.05.28.21258008

**Authors:** Meg L. Small, Robert P. Lennon, John J. Dziak, Rachel A. Smith, Gillian Sommerville, Nita Bharti

**Author notes:** **Corresponding author:**, Meg Small, Edna Bennett Pierce Prevention Research Center, 313 BBH Building, University Park, PA 16802.

## Abstract

On college campuses, effective management of vaccine-preventable transmissible pathogens requires understanding student vaccination intentions. This is necessary for developing and tailoring health messaging to maximize uptake of health information and vaccines. The current study explored students’ beliefs and attitudes about vaccines in general, and the new COVID-19 vaccines specifically. This study provides insights into effective health messaging needed to rapidly increase COVID-19 vaccination on college campuses—information that will continue to be informative in future academic years across a broad scope of pathogens. Data were collected via an online cohort survey of college students aged 18 years and older residing on or near the campus of a large public university during fall 2020. Overall, we found COVID-19 vaccine hesitancy in college students correlated strongly with some concerns about vaccines in general as well as with concerns specific to COVID-19 vaccines. Taken together, these results provide further insight for message development and delivery, and can inform more effective interventions to advance critical public health outcomes on college campuses beyond the current pandemic.

## Introduction

Colleges and universities provide important experiences that support young adults’ development, including residential education, congregate living, social opportunities, and work in labs and artistic spaces. The social interactions that occur in these spaces are core to universities’ design. In addition, social interactions in retail and hospitality contribute significantly to local economies. Interactions among college students and between college students and local communities were disrupted during the COVID-19 pandemic in order to comply with public health guidelines to slow virus spread. With the development of vaccines, social interactions can safely resume when the proportion of the population immunized against SARS-CoV-2 is very high^1^. Estimates suggest around 60-70% of the population needs to be immune to interrupt local chains of transmission, though these thresholds vary with estimates of the duration of protective immunity and the structure of contact networks. The duration of immunity from natural infection or vaccination remains unknown, with evidence suggesting significant declines in antibody levels in the months following natural infection^2^. Additionally, these factors may change over time and differ by location^3,4^ indicating that maintaining high levels of population immunity will present an ongoing challenge. Achieving high levels of vaccine uptake is the most effective strategy for safely resuming campus and community activities during the current COVID-19 pandemic. COVID-19 vaccines will be a critical tool for addressing new SARS-CoV-2 variants and seasonal fluctuations for years to come.

In the U.S., multiple COVID-19 vaccines have been authorized for emergency use^5^. These vaccines are shown to be safe and effective at significantly reducing severe disease and hospitalization^6^, with increasing evidence for reducing transmission as well^7^. Identifying the predictors of college student intentions to get vaccinated is important to design tailored strategies to increase uptake and reduce the immediate COVID-19 threat. Colleges can further help ensure students move into adulthood informed and empowered to apply scientific information when making future health decisions by teaching the skills necessary to assess and apply scientific information. For instance, an understanding of student vaccine beliefs and intentions can inform innovative instruction that helps students develop critical thinking skills to understand issues specific to their concerns or interests and encourage them to use scientific information when making health decisions.

While vaccine intentions fluctuate, current evidence suggests that vaccine hesitancy remains a challenge. A large international study of nursing students revealed that only 43% intended to get vaccinated^8^. Predictors included being male, receiving the flu vaccine, and trust in the healthcare system. A recent study of adults living adjacent to a large university found that trust in the vaccine development system and potential side effects predicted COVID-19 vaccine hesitation^9^. However, according to the Pew Research Center, adults are increasingly willing to get vaccinated with racial and gender differences narrowing and political party influence increasing^10^.

Despite the abundance of research on the predictors of COVID-19 vaccination intentions and behavior among adults, research involving college students is limited. College students represent over 10% of the American adult population and are a specific subset of adults who reside in high density housing with a large number of contacts. Immunizing college students is a critical part of achieving high levels of population immunity. Along those lines, COVID-19 vaccines are unique and warrant specialized studies: although authorized for safety and efficacy, these vaccines moved through development, production, and delivery at unprecedented speed. Two of the vaccines currently being distributed rely on a new vaccine approach (mRNA). As a result, reasons for COVID-19 hesitancy and suspicion may include novel factors that were not important in the uptake of other vaccines.

For U.S. college students, accepting the COVID-19 vaccine is likely situated in a complex set of factors regarding vaccines in general and specific concerns about the COVID-19 vaccine, as the pandemic has caused significant disruptions to academic life^11^. Based on pre-COVID-19 research on adult vaccination, potential predictors of getting vaccinated include attitudes and beliefs about vaccines in general, past vaccine behavior, trust in the system, and beliefs about disease risk, including susceptibility and severity^12^. Additionally, COVID-19-specific predictors and concerns may include political views and beliefs about the vaccine development process^13, 14^. The current study explored beliefs and attitudes about vaccines in general, COVID-19-specific vaccine concerns, perceptions of disease risk, perceptions and experiences of disease severity, and political views. The study also explored how these variables were correlated with each other and how concerns about vaccines in general overlapped with concerns about COVID-19 vaccines. A better understanding of these correlations will inform more effective, tailored health messaging, by targeting clusters of concerns specific to the college student population’^15 16^.

## Methods

### Sample

The current study used data from time one (T1) of a longitudinal cohort survey of college students residing on or near the campus of a large public university during fall 2020. Data were collected online between October and December 2020, prior to vaccine availability. Participants were recruited by email invitation through the university—eligibility requirements included being aged 18 years or older and enrolled full or part time during the fall 2020 semester. This study was approved by the university institutional review board. Of the 1427 who provided informed consent, 884 were able to enroll during the study’s timeframe. Current analyses included data from 696 young adult-aged students (18-29 years).

Demographic information was collected using the following items. Race was asked as a ‘select all that apply’ (*White, Black or African American, American Indian or Alaska Native, Asian, Native Hawaiian or other Pacific Islander, Some other race/write in*); Latinx (*yes, no*); and Gender (*female, male, gender nonconforming, trans male, trans female, different identity/write in*). Participants were 86% White; 11.7% Asian/Pacific Islander; 4.2% Black/African American; 7.8% Latinx; and 65% identifying as female. Participant semester standing was also collected; 32% were 1^st^ or 2nd year, 64% 3^rd^ or 4^th^ year, and 4% beyond 4^th^ year. The overall university undergraduate population is 76.5% White, 4.8% African American, 13.2% Asian American/Pacific Islander, and 8.4% Hispanic/Latino; and 42.0% identifying as female. Similar to our sample, the overall university undergraduate population is 48.0 1^st^ or 2^nd^ year and 61.0% 3^rd^ or 4^th^ year.

### Measures

The outcome variable was COVID-19 vaccination intention. Participants were asked, “If an FDA-approved vaccine to prevent COVID-19 were available today at no cost to you, how likely would you be to get vaccinated?*”*, answering on a 5-point scale (1=*very unlikely*, 5=*very likely*). Attitudes and beliefs about vaccines in general included side effects, trust, and cynicism. Potential side effects, measured by asking participants if they worried about unknown side-effects and unforeseen problems in children (1=*strongly disagree*, 5=*strongly agree*). Trust was measured as whether the respondent trusted the current system for evaluating vaccine safety (1 item, 1=*strongly disagree*, 5=*strongly agree; M=3*.*5*), and vaccine cynicism (two items, *r* = .61; e.g., M=1.5, e.g., *Authorities promote vaccination for financial gain, not for people’s health; vaccination programs are a big con*). Specific concerns about the COVID-19 vaccines were measured by checking all that applied *(Not sufficiently evaluated for safety; Not sufficiently evaluated for efficacy; Political pressure interfered with the regulatory process; Not enough minority participants were enrolled in studies; It will take too long for people like me to have access)*. COVID-19 susceptibility was measured with one item *My chance of getting COVID-19 is high* (1=*strongly disagree*, 5=*strongly agree*). For students who reported having a positive COVID-19 test result, severity was measured with one item *How negatively was your physical health affected (1=not at all, 5=very negatively);* for those who did not report having a positive COVID-19 test, perceived severity was measured with one item *If you were to be infected with COVID-19 how negatively do you think your health would be affected (1=not at all, 5=very negatively)*. Political viewpoint was measured with one item *Generally speaking how do you view your own political beliefs* (1=*very conservative*, 7=*very liberal*). Past behavior was measured by asking participants whether they had received a flu vaccine since September 1, 2019 (*yes, no, I wanted to get the flu vaccine, but was unable to for medical reasons*).

## Data Analyses

Pearson correlations were calculated between pairs of variables, treating the ordinal variables as numeric for ease of interpretation. Missing values were excluded pairwise. The variables used in the analysis had very low amounts of missingness (<1%). Exceptions were the political conservatism item (4% of respondents chose not to answer) and the illness severity item, which was coded as two separate variables—one for those who had tested positive (actual severity) and one for those who had not tested positive for COVID (perceived severity).

## Results

### Factors correlated with vaccine hesitancy

We identified five factors for which student responses were strongly positively correlated with COVID-19 vaccine hesitancy (correlation range (0.28, 0.40)) (Fig. 1, top rows). Starting with the factor most strongly positively correlated with vaccine hesitancy, the top five were: worries about unknown side-effects, worries about unforeseen problems in children, vaccine cynicism, worries that the COVID-19 vaccine was not sufficiently evaluated for safety, and political conservatism. Of these top five factors, three were concerns about vaccines in general, one was specific to COVID-19 vaccines, and one was an individual characteristic.

**Figure 1.**
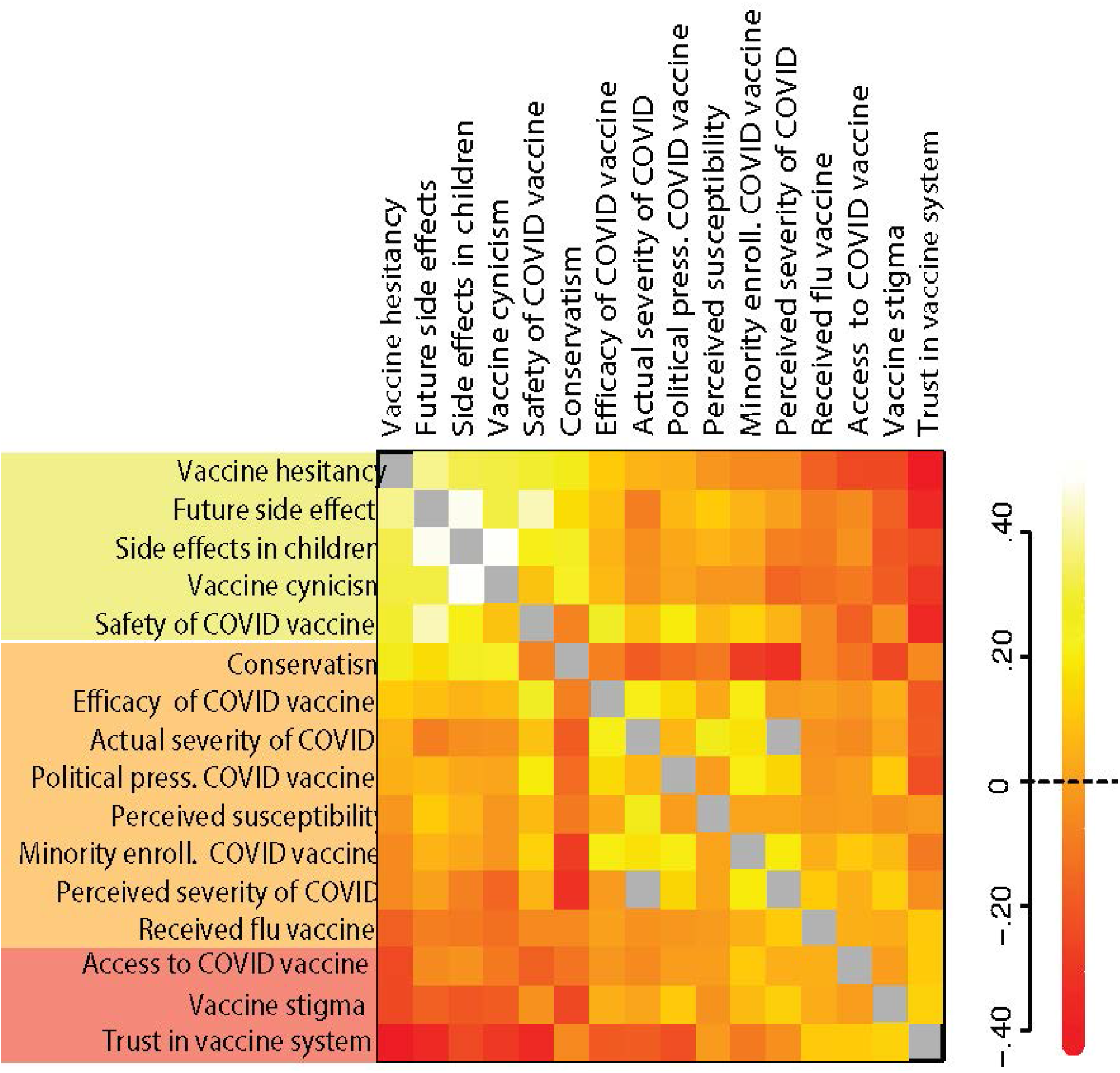
Correlation Matrix. Pairwise correlations of all the factors studied with student survey responses. The first column and first row show COVID-19 vaccine hesitancy. White and bright yellow represent the strongest positive correlational values dark reds represent the strongest negative correlational values. The diagonal of self-correlation is shown in grey, NA values are shown in grey.

We identified three factors that were strongly negatively correlated with COVID-19 vaccine hesitancy (correlation range −0.42, −0.23); (Fig. 1, bottom rows). Starting with the factor most strongly negatively correlated with vaccine hesitancy (or positively correlated with vaccine acceptance), these were: trust in the current system for evaluating vaccine safety (p<.0001), belief that stigma was not associated with receiving the COVID-19 vaccine (p<.0001), and a belief that it would not take too long for people like them to have access to a COVID-19 vaccine (p<.0001). Of these three, one was a concern about vaccines in general and two were specific to COVID-19 vaccines.

Of the remaining seven factors we evaluated, three showed weak positive correlations with COVID-19 vaccine hesitancy (correlation range 0.06, 0.13); concerns about the efficacy of COVID-19 vaccines (p=.001) and the perceived severity of COVID-19 if infected (p<0.26); and concerns that political pressure interfered with the regulatory process for COVID-19 vaccines (p<0.14). Three showed weak negative correlations with vaccine hesitancy (correlation range −0.04, −0.16): having previously received the influenza vaccine (p<.0001), the extent to which physical health had been affected by those reporting a prior COVID-19 infection (p=0.48), and concerns about minority enrollment in COVID-19 vaccine studies (p=0.36). One showed no correlation with vaccine hesitancy (correlation (−0.01): a perceived high susceptibility to COVID-19) (Fig. 1, middle rows).

### Correlations between factors

We found high correlations between responses among some factors. Factors for which responses were most strongly positively correlated with COVID-19 vaccine hesitancy were also strongly correlated with each other. Students responded very similarly to concerns about vaccines in general regarding future side effects, concerns about side effects in children, vaccine cynicism, and specific safety concerns about COVID-19 vaccines.

Responses on conservatism were strongly positively correlated with concerns about vaccines in general causing side effects in children and vaccine cynicism; these factors were strongly positively correlated with COVID-19 vaccine hesitancy as well. We also found a high correlation between responses relating to concerns about vaccines in general, specifically regarding safety and about COVID-19 vaccines regarding efficacy. Concerns about vaccine safety were strongly positively correlated with COVID-19 vaccine hesitancy but concerns about COVID-19 vaccine efficacy were only moderately positively correlated with COVID-19 vaccine hesitancy. For students who had previously been infected, their beliefs about their own chances of getting COVID-19 were strongly correlated with the severity of their COVID infection.

While the current study did not explore racial or gender differences as predictors of hesitancy, there were significant demographic differences in *specific concerns* about the COVID-19 vaccines. Female students were more concerned than male students about insufficient safety evaluation (64% vs. 55%, chi-sq=4.82, p<.05) and about minority enrollment (24% vs. 15%, chi-sq = 6.33, p=.01). However, female students were less concerned than male students about political pressure (45% vs. 59%, chi-sq=11.52, p<.0001). Black and African American students were more concerned than non-Black students about the efficacy of the COVID-19 vaccine (59% vs. 34%, chi-sq = 6.63, p=.01) and minority enrollment (55% vs. 20%, chi-sq=19.0, p<.0001).

### Limitations

This study had several limitations. The non-probability sample may not have represented the overall student population, with greater representation among White and female students. The single timepoint prior to vaccine availability limited the predictors to those based on other vaccines and evolving information about COVID-19.

## Discussion

Approaching high levels of population immunity by fall 2021 is important to colleges and universities hoping to reopen and welcome students back to pre-pandemic activities in campus classrooms, dorms, and event venues. High vaccine coverage is important to protect students, faculty, staff, and the residents of college town communities. This coverage is only achievable with strong, targeted messaging to reach those who are vaccine hesitant. While concerns about vaccines *in general* can help inform communication and intervention efforts^16^, understanding how these factors cluster with COVID-19 specific concerns and impact vaccination behavior is critical^17^. Factors such as trust in the evaluation system and concerns about side effects together can inform better intervention strategies to promote vaccination. Trust and safety are broad concepts: an understanding of the aspects of trust and safety more salient for students is needed to better target and tailor messaging. Similarly, individual factors such as political views provide insight relevant to the current COVID-19 vaccine and therefore are important considerations for message development and delivery. Predictors of vaccine hesitancy are likely to change over time^18^ and thus ongoing research will be needed to detect emerging issues. Tailoring messages to address the factors most likely to impact specific college students’ beliefs and behaviors can increase vaccination rates in the near term, and may also help build a foundation of trust in the scientific process. This study provides insight for both researchers and practitioners responsible for campus health to better create and target messaging specifically for college students. Additionally, college represents a developmental stage and an important setting for learning to critically evaluate scientific information when making health and vaccine decisions. The current vaccination context provides a potentially high return on investment for immediate and future public health.

## Data Availability

Direct data queries to the corresponding author

## Acknowledgments

The authors thank the members of the D4A Action Research Group: Dee Bagshaw, Clinical & Translational Science Institute; Cyndi Flanagan, Clinical Research Center; Matthew Ferrari, Dept. of Biology & Huck Institutes of the Life Sciences; Thomas Gates, Social Science Research Institute; Margeaux Gray, Dept. of Biobehavioral Health; Suresh Kuchipudi, Animal Diagnostic Lab; Vivek Kapur, Dept. of Animal Science and the Huck Institutes of the Life Sciences; Stephanie Lanza, Dept. of Biobehavioral Health and the Prevention Research Center; James Marden, Dept. of Biology & Huck Institutes of the Life Sciences; Susan McHale, Dept. of Human Development and Family Studies and the Social Science Research Institute; Glenda Palmer, Social Science Research Institute; Andrew Read, Depts. of Biology and Entomology, and the Huck Institutes of the Life Sciences; Connie Rogers, Dept. of Nutritional Sciences and the Huck Institutes of the Life Sciences; and Charima Young, Penn State Office of Government and Community Relations.

## Funding Source

This work was supported by the Office of the Provost and the Clinical and Translational Science Institute, the Huck Institutes of the Life Sciences, and the Social Science Research Institute at the Pennsylvania State University.

## Conflicts

The authors declare that there is no conflict of interest.

## IRB approval

This project was approved by the Pennsylvania State University IRB, #00015547.

## Notes

### Competing Interest Statement

The authors have declared no competing interest.

### Author Declarations

This study was approved by the Penn State Institutional Review Board

